# Confinement or internment? Aeromedical retrieval of pregnant people in labour: A retrospective observational study

**DOI:** 10.1101/2022.10.27.22281631

**Authors:** Bridget Honan, Breeanna Spring, Fergus William Gardiner, Cheryl Durup, Ajay Venkatesh, Jessica McInnes, Rebecca Schultz, Shahid Ullah, Richard Johnson

**Affiliations:** Central Australian Retrieval Service, Alice Springs, NT, Australia; Royal Flying Doctor Service of Australia, Canberra, ACT, Australia; Molly Wardaguga Research Centre, College of Nursing and Midwifery, Charles Darwin University, Darwin, NT, Australia; University of Western Australia, Crawley, WA, Australia; Alice Springs Hospital, Alice Springs, NT, Australia; School of Medicine and Dentistry, Griffith University, Southport, QLD, Australia; Edith Cowan University, Joondalup, WA, Australia; School of Medicine and Public Health, Flinders University, Bedford Park, SA, Australia; Baker Institute, Melbourne, VIC, Australia

**Author notes:** Corresponding author: Dr Bridget Honan.

**Keywords:** Air Ambulances, Rural Health Services, Obstetric Labour, Premature

## Abstract

**Introduction:** The aim of this study was to describe the characteristics and outcomes of remote-dwelling pregnant people with threatened labour referred for aeromedical retrieval to a regional birthing centre, as well as factors associated with birth within 48 hours.

**Methods:** This was a retrospective observational study of all pregnant people in the remote Central Australian region referred to the Medical Retrieval Consultation and Coordination Centre for labour >23 weeks gestation, between 12 February 2018 – 12 February 2020. Data was extracted manually from written medical records on maternal, neonatal and retrieval mission characteristics. Univariate and multivariate statistical analysis was performed.

**Results:** There were 116 people referred for retrieval for labour. There were no births during transport and less than half of the cases in this cohort resulted in birth within 48 hours of retrieval. Tocolysis was frequently used. Predictors of birth with 48 hours were cervical dilatation 5cm or more, preterm gestational age and ruptured membranes in the univariate analysis. Nearly one-third of this cohort required intervention or had complications during birth.

**Discussion:** Birth during transport for threatened labour did not occur in this cohort, and more than half of retrievals did not result in birth within 48 hours, however the high risk of birth complications may offset any benefit of avoiding aeromedical transport from remote regions. Retrieval clinicians should have a lower threshold for urgent transfer in cases of ruptured membranes, cervical dilatation of 5cm or more, or gestational age is less than 37 weeks.

## Introduction

Pregnant people living in rural and remote areas of Australia have less access to maternity services than those living in metropolitan areas (1,2) have higher rates of obstetric complications including premature birth (1,3–7), and are reliant on aeromedical retrieval services for unplanned care (2,8,9). Despite preterm labour being one of the most common reasons for aeromedical retrieval in Australia, and poorer health status for rural and remote Australians, there is little published literature on maternal and infant outcomes following aeromedical retrieval for labour (10).

Birthing services in Australia are designed around regionalised models of care, with specialised services located vast distances from remote communities. Specialised care reduces maternal and infant morbidity and mortality, such as stillbirth and neonatal death, particularly amongst mothers and babies with risk of complications and prematurity (11). However, this model of care displaces pregnant people from their local communities, families, customs, and traditional birthing practices, which detrimentally impacts on cultural safety and wellbeing (12,13). For this reason, it is imperative to avoid unnecessary transfers.

Understanding the trajectory of labour and predicting time-to-birth is important to inform decision-making about timing of retrieval and other treatment options including tocolysis and steroids for foetal lung maturation (14). Furthermore, decisions about the timing of aeromedical evacuation of a pregnant person in suspected labour involve prioritising limited retrieval resources whilst avoiding birth during transport. Knowledge of the incidence of birth complications in this cohort, including post-partum haemorrhage and surgical delivery, is also important in planning for obstetric emergencies and services in rural and remote areas.

A recent scoping review found that birth during aeromedical transport of pregnant women in labour from remote areas is very rare, but little is known about predictors of time-to-birth, and there is a need for data on this topic to understand the needs of pregnant people with threatened labour located remotely from a birthing centre (10).

The primary objective of this study is to describe the maternal and infant characteristics and outcomes for people referred to the retrieval service for labour in a geographically remote area. The secondary objectives are to report time-to-birth and associated factors.

## Methods

### Setting

This study was conducted in Central Australia, a region of 1.6 million square kilometres and a population of 60,000 people, of which more than half live in remote communities. The catchment area encompasses Alice Springs, Barkly and APY lands areas as defined by the Australian Bureau of Statistics (15). The Alice Springs Hospital is the regional hospital with 180 beds, including maternity and paediatric services and an Intensive Care Unit, but no specialist neonatal services.(16)

The medical facilities in Central Australian remote communities consist of an outpatient clinic and an emergency room, without any capacity to admit inpatients. Staffing in remote community clinics is Remote Area Nurse led, with the largest communities having resident or visiting primary care doctors and/or midwives. None of the referring centres are designated as birthing facilities and do not provide maternity services. Local policy requires pregnant people to travel to Alice Springs prior to birth to access maternity services (8,17).

The Medical Retrieval and Consultation Centre (MRaCC) is staffed by critical care medical specialists who coordinate the retrieval response to medical emergencies in the Central Australian region. The details of MRaCC can be found elsewhere (18). The service also coordinates transfers of patients from the Alice Springs hospital to the quaternary hospitals located interstate, such as the Royal Adelaide Hospital.

The retrieval service utilises fixed-wing aircraft and crew available over 4 shifts in a 24-hour period provided by the Royal Flying Doctor Service (RFDS)(19). The crew may consist of Flight Nurses (dual trained Registered Nurse and Registered Midwives) and Retrieval Doctors (who are trained in critical care specialities such as Emergency Medicine however may have limited skills in obstetric or neonatal care). A paediatrician may join the retrieval team if the risk of birth is thought to be significant.

### Participants

All pregnant people with an estimated gestational age of >23 weeks who were referred to MRaCC for retrieval due to labour at the time of referral, were eligible for inclusion. The criteria for labour are necessarily subjective, because in most cases, the assessment and referral are performed by someone without expertise in midwifery. Typical symptoms of labour include onset of regular painful contractions.

Participants were included if they required a retrieval from 12 February 2019 – 12 February 2020. This period was chosen because it coincides with the commencement of the MRaCC model of retrieval coordination in the region, and avoids potential anomalies associated with onset of the COVID-19 pandemic.

Participants were excluded from analysis if the reason for referral for retrieval was rupture of membranes or vaginal bleeding without signs or symptoms of labour, or if they were referred for interhospital transfer to a specialist neonatal facility, without any signs of symptoms of labour. Participants were also excluded if they were referred for foetal death-in-utero or not thought to be of viable gestational age.

### Outcomes

Time-to-birth was defined as the time interval between onset of signs or symptoms of labour (such as strong regular contractions), and the time of birth, and categorised as occurring either within 48 hours or more than 48 hours after onset of labour. This cut-point was chosen because birth may be delayed by medical interventions for 48 hours to optimise neonatal condition (for example, steroids for foetal lung maturation). Births after 48 hours suggest that the diagnosis of threatened labour may not have been accurate (20). Outcomes are shown in Table 1.

**Table 1.** Participant characteristics and outcomes

| **Characteristics** | **N (%)** |
| --- | --- |
| ***Transfer characteristics*** |  |
| Retrievals | 116 |
| Suspected preterm labour (<37 weeks gestation) | 85 (73.9%) |
| Time from referral to first retrieval contact, h, Median (IQR) | 3.3 (2.8 - 4.8) |
| Time from referral to arrival time at maternity hospital, h, Median (IQR) | 5.7 (4.5-7.6) |
| > 1 retrieval during same pregnancy | 8 (6.9%) |
| ***Maternal characteristics at time of retrieval*** |  |
| Median mother age, y, Median (IQR) | 25 (21.8-30) |
| Nulliparous (parity = 0) | 37 (31.9%) |
| Multiparous (parity >=1) | 79 (68.1%) |
| BMI >35 at time of birth, kg/m<sup>2</sup> | 23 (22.5%) |
| Diabetes | 16 (13.8%) |
| Hypertension | 9 (7.8%) |
| Rheumatic heart disease confirmed or suspected | 11 (9.5%) |
| ***Maternal assessment during prehospital encounter*** |  |
| Gestational age <37 weeks at time of referral | 85 (73.9%) |
| Suspected uterine infection (defined as any of: documented fever >38C, uterine tenderness or purulent vaginal discharge). | 21 (18.1%) |
| Ruptured membranes | 46 (39.7%) |
| Cervical examination performed | 69 (59.5%) |
| Cervical dilatation <5cm (if examined) | 58 (84.1%) |
| Cervical dilatation >=5cm (if examined) | 11 (15.9%) |
| ***Treatments given prehospital or during retrieval*** |  |
| Tocolysis given prehospital or during transport | 91 (78.4%) |
| Nifedipine given prehospital or during transport | 90 (98.9%) |
| Magnesium given prehospital or during transport | 1 (1.1%) |
| Tocolysis given when estimated gestational age 35+ weeks | 37 (32.2%) |
| Tocolysis given in presence of suspected uterine infection | 17 (14.8%) |
| Requiring a blood transfusion during retrieval | 2 (1.7%) |
| Magnesium with gestational age <30 weeks at time of referral | 0 |
| Steroids given with EGA <35 weeks at time of referral | 41 (35.7%) |
| Antibiotics given | 67 (57.8%) |
| Opioid analgesia given | 20 (17.5%) |
| ***Crew mix*** |  |
| Retrieval doctor accompanied mission | 101 (87.1%) |
| Pediatrician accompanied mission | 4 (3.4%) |
| ***Birth characteristics and outcomes*** |  |
| Time-to-birth from onset of labour, h, Median (IQR) | 64 (17.5 – 618.6) |
| Birth within 48 hours of onset of labour | 46 (45.1%) |
| No details about birth available | 4 (3.4%) |
| ***Birth location*** |  |
| Hospital | 98 (87.5%) |
| Clinic or hospital without maternity services | 13 (11.6%) |
| Home | 1 (0.9%) |
| Retrieval platform | 0 |
| ***Birth complications*** |  |
| Instrumental vaginal delivery | 8 (7.1%) |
| Lower uterine Segment Caesarean Section | 27 (24.1%) |
| PPH occurred | 33 (29.5%) |
| ***Premature birth*** |  |
| Born at premature gestation (<37 weeks) | 43 (38.4%) |

Note. Number and percentages are reported unless stated otherwise; IQR Interquartile range

### Data sources

Data were manually extracted from electronic and paper medical records by two retrieval doctors (CD and AV) into a spreadsheet. The following sources were used: retrieval service databases (for information pertaining to retrieval service delivery including time of referral and use of medications); primary care medical records (for information on antenatal care, maternal characteristics, and initial assessment of threatened labour) and hospital medical records (for information on obstetric care and birth outcomes).

### Statistical analysis

All statistical analyses were performed using Stata statistical software, version 16.1. Participant’s characteristics at birth were expressed as absolute and relative frequencies for categorical variables and were compared between exposure groups using a standard Chi-square test for association with continuity correction, where appropriate. Mean and standard deviation (SD) for continuous data were also be presented and an independent sample t test was used to explore the significant differences of patients’ characteristics between groups. Data normality was visually checked using frequency histograms and normal Q-Q plot. For non-normally distributed data, median and interquartile ranges (IQR) was also be reported. A logistic regression model was applied to examine the time to birth outcome between exposure groups. The time to birth was categorised into <=48 hours and >48 hours measured from the time of onset of labour to the time of birth.

Univariate models were first performed without adjustment. Then, multivariate analysis was undertaken, by adding covariates considered clinically important and statistically significant from the univariate comparisons, to adjust for confounding between variables. The estimates were calculated using the maximum likelihood method and were expressed as odds ratios (ORs).

Model diagnostics and goodness of fit were evaluated by Receiver Operator Characteristic (ROC) curves. The two-sided test was performed for all analysis and the level of significance was set at p < 0.05.

## Results

There were 5245 referrals to MRACC during the study period, of which 183 were categorised as obstetric-related retrievals. Of these, 116 (63.4%) referrals were for in labour. 4 cases were excluded due to missing time and location of birth.

Participant characteristics and outcomes are shown in Table 1.

Retrieval mission time from referral to arrival at hospital was a median of 5.7 hours (IQR 4.5-7.6). In terms of interventions, cervical examination was performed in 59.5% of cases, and tocolysis was given in 78.4% of cases.

73.9% of participants were of preterm gestational age at the time of retrieval, although 38.4% infants were premature at time of birth. The median time-to-birth was 64 hours (IQR 18 – 619 hours). Overall, 46 participants (45%) delivered within 48 hours of onset of labour. There were 4 births before hospital.

Complications or interventions during birth included instrumentation in 7.1% of births, caesarean section in 24.1% of births and PPH in 29.5% of births. 2 participants required blood transfusion during transport. There were no maternal deaths.

In the univariate analysis, factors that were significantly associated with birth within 48 hours were premature gestational age at time of referral (<37 weeks), ruptured membranes or cervical dilatation of 5cm or more (Table 2). In the multivariate analysis, only ruptured membranes remained significantly associated with birth within 48 hours (Table 3). Cervical examination was not included due to no cases of cervical dilatation >5cm birthing after 48 hours.

**Table 2.**
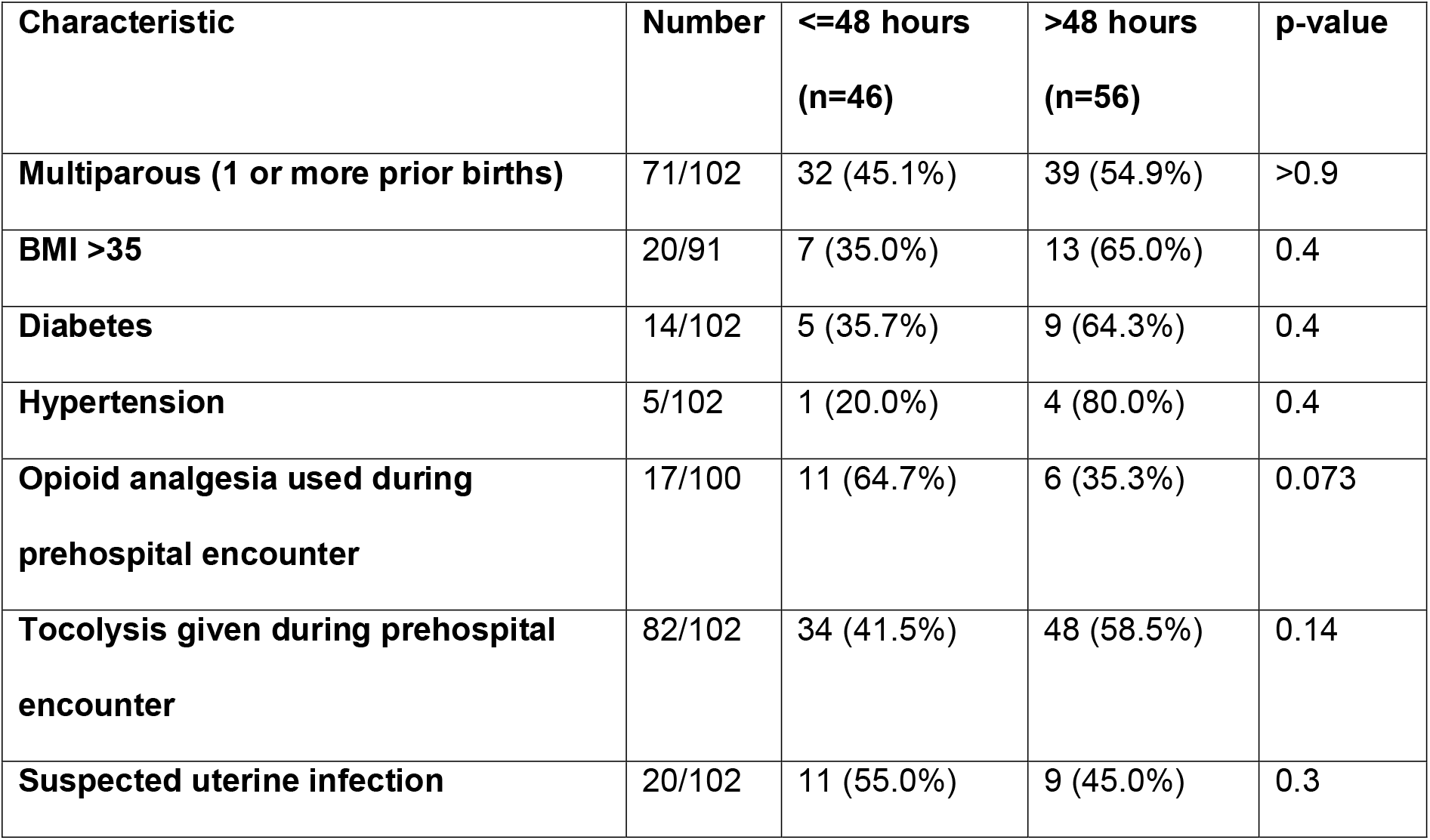
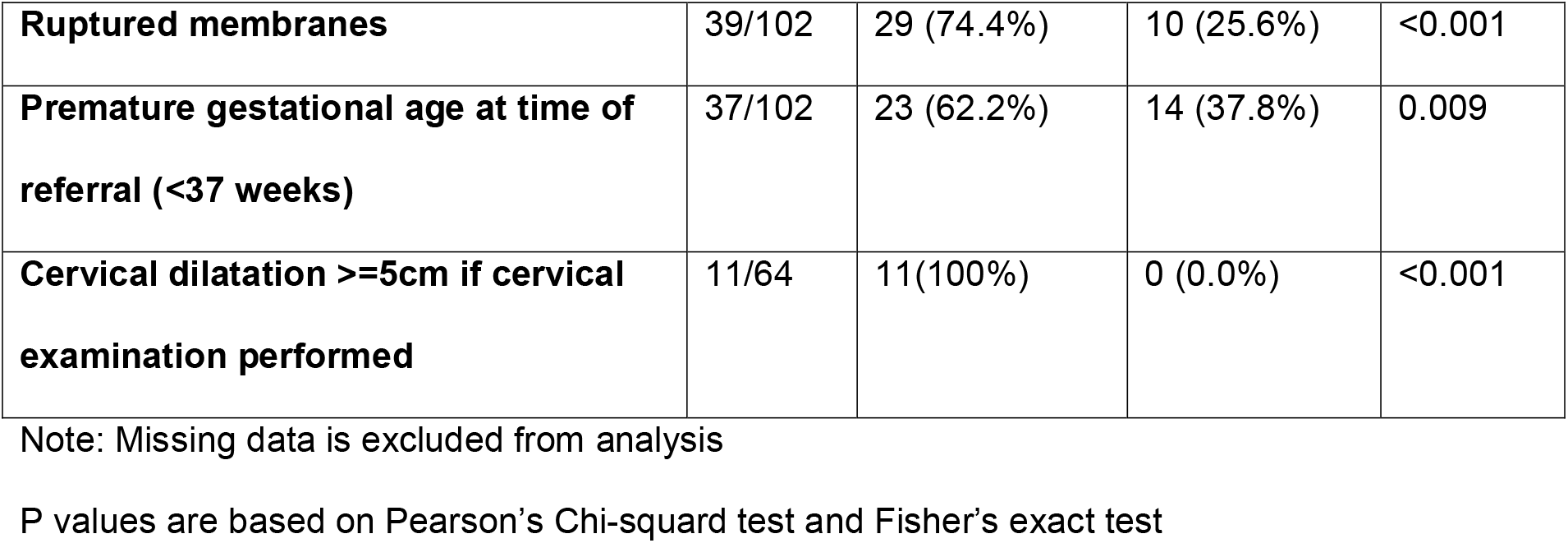
Association between birth within 48 hours and other variables using univariate logistic regression analysis.

| Characteristic | Number | <=48 hours<br>(n=46) | >48 hours<br>(n=56) | p-value |
| --- | --- | --- | --- | --- |
| **Multiparous (1 or more prior births)** | 71/102 | 32 (45.1%) | 39 (54.9%) | >0.9 |
| **BMI >35** | 20/91 | 7 (35.0%) | 13 (65.0%) | 0.4 |
| **Diabetes** | 14/102 | 5 (35.7%) | 9 (64.3%) | 0.4 |
| **Hypertension** | 5/102 | 1 (20.0%) | 4 (80.0%) | 0.4 |
| **Opioid analgesia used during prehospital encounter** | 17/100 | 11 (64.7%) | 6 (35.3%) | 0.073 |
| **Tocolysis given during prehospital encounter** | 82/102 | 34 (41.5%) | 48 (58.5%) | 0.14 |
| **Suspected uterine infection** | 20/102 | 11 (55.0%) | 9 (45.0%) | 0.3 |
| **Ruptured membranes** | 39/102 | 29 (74.4%) | 10 (25.6%) | <0.001 |
| **Premature gestational age at time of referral (<37 weeks)** | 37/102 | 23 (62.2%) | 14 (37.8%) | 0.009 |
| **Cervical dilatation ≥5cm if cervical examination performed** | 11/64 | 11(100%) | 0 (0.0%) | <0.001 |

Note: Missing data is excluded from analysis
P values are based on Pearson's Chi-squared test and Fisher's exact test

**Table 3.**
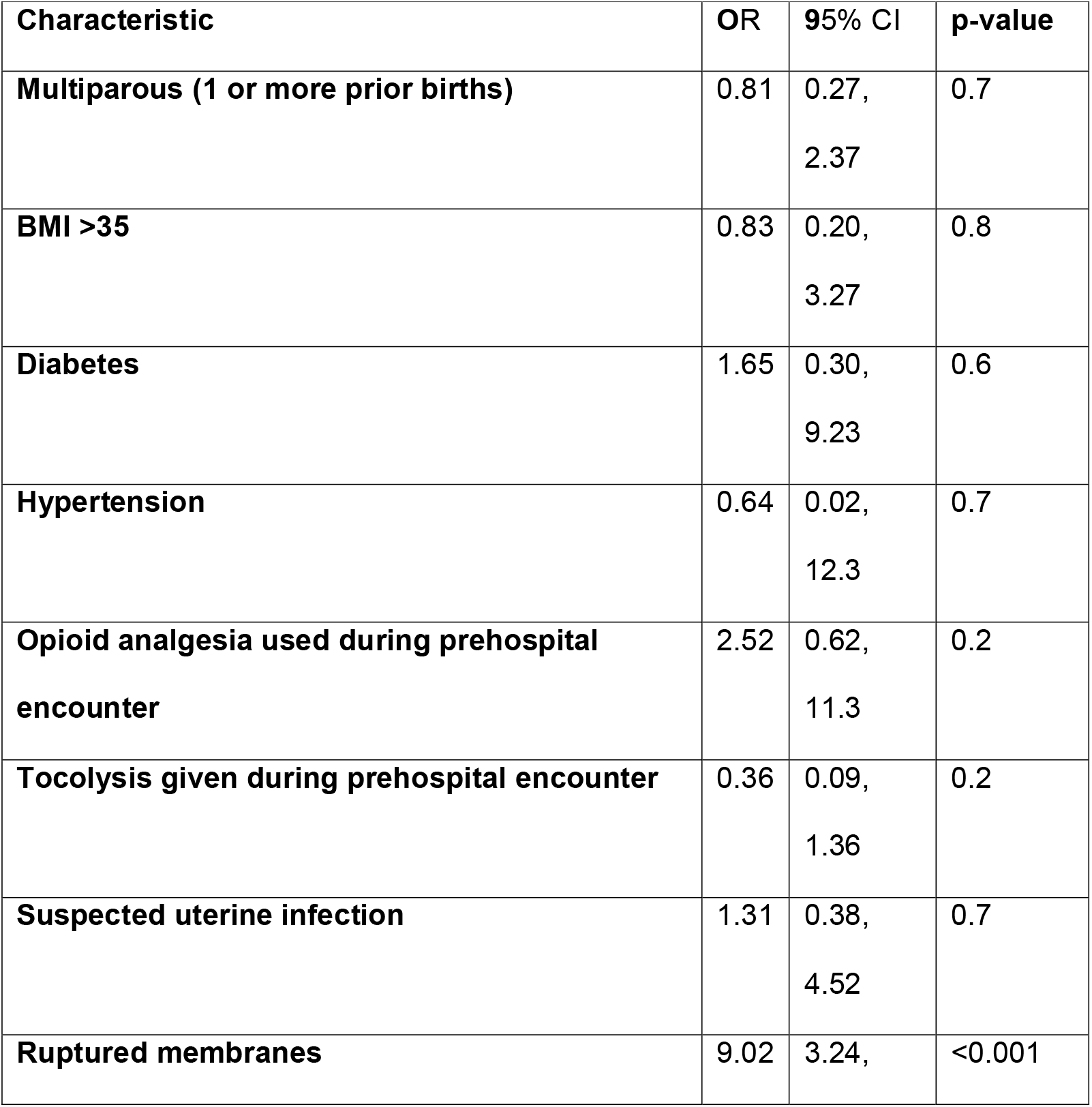
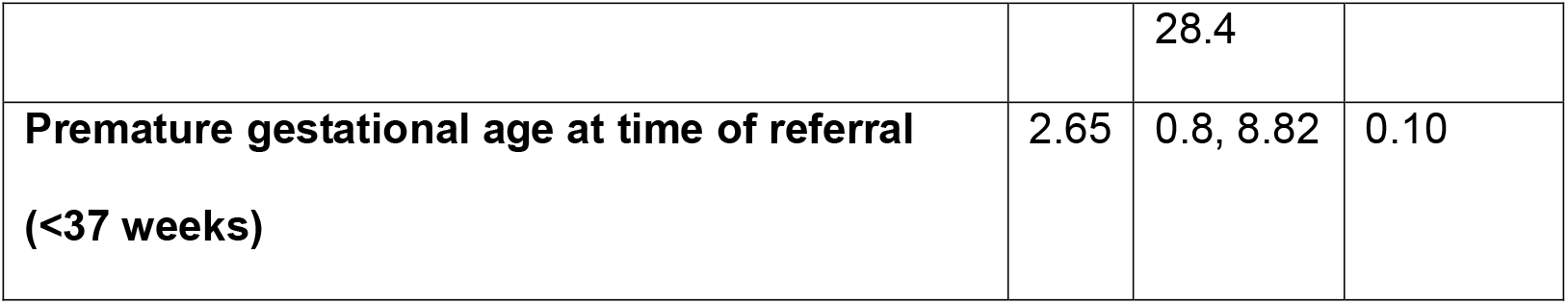
Multivariate logistic regression model of time to birth <=48 hours for maternal characteristics at birth

## Discussion

In this retrospective observational study of pregnant people in labour referred to an aeromedical retrieval service for transport to a birthing centre, there were no births during transport despite long transport times, and more than half did not deliver within 48 hours of onset of symptoms of labour. However, the high level of maternal comorbidity and birth complications in this cohort are important considerations for remote maternity and retrieval services.

Previous studies on aeromedical retrieval for people in labour suggest that birth during transport is rare (21,22). One study from Western Australia found that no people transported for imminent risk of preterm birth delivered during the aeromedical transport (23). Similarly, another study in the top end of the Northern Territory of Australia found that there were 5 births in the remote clinic, 1 at the remote airstrip and 1 in the ambulance between the clinic and the destination hospital, but no births in-flight (24). In metropolitan retrieval services, retrieval physicians are rarely involved in birth or obstetric interventions (21,25).

Even with specialist assessment, predicting the trajectory of labour can be inaccurate. Some studies have shown that less than 10% of people with a clinical diagnosis of preterm labour will birth within 7 days of initial presentation (26,27), and almost 75% will birth at term (28). In this cohort, 49% of retrievals for threatened preterm labour ultimately birthed at term.

Pre-hospital and retrieval medicine experts recommend that a patient with cervical dilatation greater than 5cm, should not be transferred due to the risk of precipitous birth in-flight(14). One study of rural women transferred to a tertiary hospital for signs of preterm labour showed that cervical dilatation (in conjunction with nulliparity and regular contractions) was associated with shorter time-to-birth(29). In nearly half of all cases in this cohort, no cervical examination was performed, possibly due to lack of confidence in performing the exam, or because the examination would not change the need for retrieval. In cases where cervical examination was performed, all women who had cervical dilatation >5cm delivered within 48 hours.

In our univariate analysis, premature gestational age or ruptured membranes were significantly associated with birth within 48 hours in the univariate analysis. However, in the multivariate analysis, only ruptured membranes remained significantly associated with birth within 48 hours. Although there is conflicting data on whether preterm prelabour rupture of membranes is associated with shorter time-to-birth(29–32), ruptured membranes in conjunction with uterine activity has been shown to predict birth within 7 days(33).

The high proportion of continuing pregnancies after initial retrieval suggest there is a high degree of over-triage and potentially avoidable transfers. The threshold for retrieval is low because of the limited access to life-saving obstetric interventions in remote settings. This is reflected in the high rate of maternal and neonatal complications demonstrated in this cohort, with PPH occurring in 29.5% of births, and instrumental or surgical birth in 31.2% of births. This compares unfavourably to other Australian data, for example, with rate of PPH in Queensland previously reported at 6.9%(34). Similarly, a case series of 4 in-flight births over a 4-year period in the Northern Territory found a high rate of morbidity and mortality, including 1 case of APH, 2 cases of PPH, 3 cases of neonatal resuscitation and 1 neonatal death 2 hours after birth (35), illustrating the rationale for urgent transfer to avoid outborn birth.

It is therefore not surprising that tocolysis was used liberally in this cohort. We found that tocolysis was used in cases of advanced gestations, suspected uterine infection or cervical dilatation. In most tertiary hospital guidelines, tocolysis is relatively contraindicated in these situations, because the risk of prolonging the pregnancy is greater than the risk of birth. This illustrates the need for clinical practice guidelines that reflect rural and remote practice, for remote emergency clinicians (11). A clinical trial is currently investigating the risks and benefits of administering tocolysis after preterm prelabour rupture of membranes, with important implications for rural and remote maternity services (36).

The high rate of continuing pregnancy after transfer also illustrates a potentially burdensome intervention for remote-dwelling people who are removed from their community and country for no medical obstetric benefit, which may result in cultural harms (12,13). There is some evidence to suggest that use of foetal fibronectin testing could be used to predict risk of preterm birth in pregnant people presenting to remote health services, thus reducing aeromedical transfers for premature labour(32,37–39).

For retrieval services, there is insufficient data from this study to make strong recommendations about prioritisation and timing of aeromedical retrievals for remote-dwelling pregnant people in labour, although the data suggests that there should be a lower threshold for urgent transfer in cases of ruptured membranes, cervical dilatation of 5cm or more, or gestational age is less than 37 weeks.

For rural healthcare services, the findings suggest potential targets for service delivery improvement, such as access to culturally appropriate local antenatal services, to reduce the rate of unnecessary transfers and reduce obstetrical complications.

### Limitations

As this was an observational study, association cannot be used to infer causation. The sample size was small and had missing data, which limits confidence in the findings and prevented controlling for confounders.

## Conclusion

Birth during transport for labour did not occur in this cohort, and more than half of retrievals did not result in birth within 48 hours, however the high risk of birth complications may offset the benefit of avoiding aeromedical transport from remote regions. Retrieval clinicians should have a lower threshold for urgent transfer in cases of ruptured membranes, cervical dilatation of 5cm or more, or gestational age is less than 37 weeks.

## Data Availability

All data produced in the present study are available upon reasonable request to the authors.

